# Whole-Exome sequencing identifies a Type 2 Diabetes candidate mutation in *PTPRF*

**DOI:** 10.1101/2020.09.11.20189399

**Authors:** Erik J.M. Toonen, Michael Kwint, Alexander Hoischen, Cees J. Tack

## Abstract

**Objectives:** Identification of genetic factors involved in Type 2 diabetes mellitus (T2DM) has been challenging. While genome-wide association studies (GWAS) have identified over 60 loci associated with T2DM, these variants only explain a small proportion of the total heritability of the disease. Whole-exome sequencing has become a powerful approach to identify genetic variants that are not captured by GWAS. We applied exome sequencing to identify causal genetic variants in a family with a 2-generation history of T2DM characterized by substantial insulin resistance, hypertension and isolated hypertriglyceridemia. Methods: Exome sequencing was performed on genomic DNA of two affected family members. Twenty-four identified variants that were present in both family members were further tested for segregation in all other family members by Sanger sequencing. Results: Three rare missense variants, located in the genes *PTPRF, FUCA1* and *FBXO30*, were present in all affected family members but also in one unaffected family member. Protein-protein interaction analysis showed that *PTPRF* strongly interacts with several members of the insulin signaling pathway. Conclusions: Our results suggest that the variant in the *PTPRF* gene might be causal for an unusual T2DM subtype that is characterized by a severe insulin resistance and isolated hypertriglyceridemia.

## Introduction

Type 2 diabetes mellitus (T2DM) is a complex multifactorial trait where both environmental and genetic factors contribute to the pathogenesis of the disease. Based on twin and segregation studies, the heritability of type 2 diabetes has been estimated to be between 36 and 83 percent [1, 2]. Large-scale genome-wide association studies (GWAS) have contributed towards understanding the inherited basis of diabetes. In the past few years, GWA studies have identified over numerous loci associated with T2DM [3-7]. However, these identified genetic variants account for less than 6 percent of the heritability estimated from family studies [8]. Because these common variants explain only a small fraction of the estimated heritability, it has been hypothesized that rare genetic variants with large effect sizes contribute to the genetic predisposition of T2DM. Traditional genotyping platforms used in GWAS are primarily designed to capture common genetic polymorphisms but offer poor coverage of rare genetic variants, hence rare disease variants with large effect sizes are easily missed [9]. Approaches other than GWAS are necessary to identify the remaining genetic variants underlying the pathogenesis of type 2 diabetes in order to improve diagnosis and treatment strategies.

Recently, whole-exome sequencing has become a powerful strategy to identify low-frequency and rare variants not captured by GWAS [10]. Sequencing of all genes (exome) is an approach relying on the hypothesis that functional disease-associated variation resides in the coding regions. Whole-exome sequencing has proven to be valuable for identifying mutations responsible for monogenic diseases [11] and emerging reports show that exome sequencing can also be applied to uncover variation associated with complex human traits [12, 13]. While T2DM consists of both decreased insulin secretion and reduced insulin action (insulin resistance), most genetic defects identified so far relate to insulin secretion and genetic factors in obesity-induced insulin resistance seem to be underreported [14].

In the present study, we apply exome sequencing to identify genetic variants that may have a role in disease susceptibility in a family with a 2-generation history of type 2 diabetes, with an unusual phenotype characterized by pronounced insulin resistance and hypertriglyceridemia.

## Methods

### Family members

We recruited a family with a 2-generation family history of T2DM. The index cases were under chronic diabetes care in the hospital. Due to privacy reasons, the pedigree is not published, please contact the corresponding author for further information.

Case presentations

The index case presented with diabetes in his late thirties with a six months period of weight loss (14 kg) and balanitis. Body weight at presentation 80.5 kg, height 1.73 (BMI 26.7 kg/m^2^), blood pressure 150/100. Laboratory analysis revealed HbA_1c_ 12.5%, ALAT slightly elevated, urine ketones negative. Patient started oral combination therapy (SU and metformin), with temporary improvement, but secondary glycemic failure after approximately 2 years. Before start of insulin his laboratory parameters were: HbA_1c_: 9.2%, total cholesterol 5.3, Triglycerides 4.9, HDL-cholesterol 0.70, LDL 2.51 mmol/L (all fasting and without statin), fasting insulin level: 30 mU/L, C-peptide: 1.54 nmol/L, ASAT 21, ALAT 39, urine albumin 20 mg/L. Patient was started on insulin, eventually administered by pump, up to 140 U/day (body weight increased to 94 kg, BMI 31.4 kg/m^2^). His triglyceride levels remained elevated. See Table 1 for details.

**Tabel 1.** Characteristics of family members

| Family member | Phenotype |  |  |  |  |  |  |  |  |  |
| --- | --- | --- | --- | --- | --- | --- | --- | --- | --- | --- |
|  | Diabetes | AGE of ONSET | HT | AGE | BMI (kg/m<sup>2</sup>) | Insulin dose (U/kg) | Total Cholesterol (mmol/l) | Triglycerides (mmol/l) | HDL-cholesterol (mmol/l) | LDL-Cholesterol (mmol/l) |
| 1 | Yes | 60s | Yes | 80s† | 27 | 1.9 | 4.4 - 4.7 | 3.6 – 4.7 | 0.6 | 2.2 |
| 2 | No |  | Yes | Late 80s | 35 | N/A | 4.9 | 2.5 | 0.8 | 3.0 |
| 3 | Yes<sup>#</sup> | 20s | Yes | Late 50s | 19 | 1.0 | 3.7 | 4.4 (max 25) | 0.8 | 1.0 |
| 4 | Yes | Late 30s | Yes | Late 50s | 31 | 1.6 | 5.3 | 4.9 | 0.7 | 2.5 |
| 5 | No |  | No | 50s† | ? | N/A |  |  |  |  |
| 6 | No |  | Yes | 50s | 27 | N/A | 4.8 | 1.9 | 1.1 | 2.9 |
| 7 | Yes<sup>#</sup> | Late 30s | Yes | 50s | 27 | 0.9 | 4.5 | 2.9 | 0.9 | 2.2 |
| 8 | No |  | No | Late 40s | 30 | N/A | 5.1 | 3.4 | 0.9 | 2.7 |

<sup>#</sup>denoted as type 1 diabetes but no auto-immunity tested; HT: hypertension; BMI: Body Mass Index
†Died at indicated age

A relative was diagnosed with diabetes in his sixties during routine medical examination. He was overweight (weight 87.5 kg, height 1.78, BMI 27,6 kg/m^2^) and had hypertension, micro-albuminuria, hypertriglyceridemia and low HDL-cholesterol. He was eventually treated with insulin, up to 160 U/day. He always had elevated triglyceride levels.

The clinical phenotype of affected family members was characterized by substantial insulin resistance, hypertension and hypertriglyceridemia without hypercholesterolemia. Data from one affected relative were retrieved from another hospital. Data from an unaffected relatives were obtained during a short hospital visit, data from one unaffected relative were obtained through his GP and by telephone. In family members with diabetes, detailed information regarding onset of diabetes, complications and treatment was obtained. In unaffected family members, BMI and blood pressure was measured and fasting glucose, and HbA_1c_ (to exclude diabetes) and lipids levels were measured. Results are depicted in Table 1. There was no information suggesting consanguineous marriage in the family.

### Non-related diabetes subjects

To validate our initial results obtained from the analysis of the family members, we selected 24 non-related subjects who matched the characteristics of the affected family members (insulin > 1U/kg or >100 U/day; TG > 1,5 mmol/l; BMI < 35 kg m2). Subjects were selected from the Diabetes Pearl patient cohort. This cohort is a large, well-characterized observational cohort of type 2 diabetes patients. The Diabetes Pearl cohort is a collaborative biobank initiative from the Dutch University Medical Centers (www.parelsnoer.org) [15]. DNA samples were retrieved from the Biobank after approval of the ethics committee.

### Whole-Exome sequencing

Exome sequencing was essentially performed as described previously [16, 17]. In brief, exome enrichment was performed using the SureSelect Human Exon 50-Mb Kit (Agilent, Santa Clara, CA, USA), covering approximately 22.000 genes, followed by SOLiD4 sequencing (Life Technologies, Carlsbad, CA, USA). Reads were mapped to the hg19 reference genome using SOLiD LifeScope software version 2.1. All nonsynonymous variants shared in both affected individuals and absent or with very low frequency (0.1%) in dbSNP, were further investigated in healthy and affected family members.

### Sanger Sequencing

Nonsynonymous variants identified by exome sequencing were tested in healthy and affected family members for co-segregation by traditional Sanger sequencing. Forward and reverse PCR primers for each exon in which the variant was located were selected in an intronic sequence 50 bp away from the intron/exon boundaries. Unique primers were designed using the Primer3 software. PCR products were verified on 2% agarose gel, purified using ExoSAP-IT (Affymetrix, Santa Clara, CA, USA) and analyzed on ABI3730 genetic analyzers (Life Technologies, Carlsbad, CA, USA).

Ion torrent library preparation and sequencing

All 32 coding exons of the *PTPRF* gene were analyzed in 24 unrelated diabetes patients who matched the clinical characteristics of the affected family members. The screening of 24 patients for variants in the *PTPRT* gene was carried out by using an Amplicon-based deep-sequencing approach on the Ion PGM sequencer (Life Technologies, Carlsbad, CA, USA). PCR amplicons were generated per sample using standard PCR protocols. Primer sequences are available upon request. PCR products per sample were pooled, and subsequently sheared to 200-bp fragments by the use of Covaris E210 (Covaris Inc.). Library preparation was performed using the Ion Xpress™ Plus Fragment Library Kit in combination with the Ion Xpress Barcode adapters 1-96 kit. Emulsion PCR and enrichment were carried out on the Ion OneTouch System using the Ion PGM TemplateOT2 200 Kit as described by the manufacturer. Sequencing was performed using the 200bp kitv2, with Ion Sphere particles loaded to an Ion318™ chip (Life Technologies Carlsbad, CA, USA). Raw sequencing reads were mapped to the hg19 genome using the Torrent Server with the use of the Torrent suite software. FASTQ files were subsequently analyzed and visualized in SeqNext (JSI Medical systems).

### Protein-protein interaction analysis

Protein-protein interactions (PPI) were identified using the STRING database (STRING v9.1; http://string-db.org/) [18]. STRING builds functional protein association networks based on compiled available experimental evidence. A confidence score is assigned to each identified protein-protein association and this score will increase when an association is supported by several types of evidence. In this study, only PPI are shown that yielded a high confidence score (≥0.7).

### Ethics

All family members were informed about the goal of the study and consented to participation.

## Results

### Characteristics of family members

Characteristics of affected and unaffected family members are shown in Table 1.

### Whole-Exome sequencing identifies variants within the PTPRF and ASXL1 genes

Exome sequencing was performed on genomic DNA of two family members with an average 46- and 81-fold coverage leading to 85.2% and 87.9% of the exome with at least 10-fold coverage, respectively. We identified 38,067 and 39,293 genetic variants per proband in the two family members, respectively. Variants were annotated by a bioinformatics pipeline as previously described [16]. In addition, the variant had to be present in at least 20-80% of all reads suggestive of heterozygous changes in a dominant model of disease. Subsequently, genetic variants were prioritized based on predicted amino acid consequences and overlap with common genetic variants (listed in dbSNP version 135 and/or present in an in-house database containing >10,000 analyzed exomes of predominantly European ancestry). All filter steps are shown in Supplementary Table 1.

We identified 21 unique non-synonymous variants present in both affected family members that were present in genes with known expression in adipose tissue (Table 2, Supplementary Table 1). All 21 variants were tested for segregation in all other family members by Sanger sequencing (Table 3). Two rare missense variants were present in all affected family members but also in one unaffected family member. Variants c.3614A>G (rs200885607; p.(N11965S); located in the gene *Protein tyrosine phosphatase, receptor type F (PTPRF;* NM_002840)) and c.904C>T p.(R302C); located in the gene *ASXL transcriptional regulator 1* (*ASXL1*; NM_015338)) were also present in the non-affected family member II/4 (Table 3). The c.3614A>G variant within *PTPRF* was detected once in the in genome-wide sequence data from the 1000 Genomes Project which include the genomes of 1088 people (or 2176 chromosomes) [19]. The variant was present at very low frequency in public exome datasets, e.g. ExAc browser frequency 12/121398 alleles, i.e. allele frequency of 9.885 x 10^-^5; with 7/66730 alleles in Non-Finish Europeans (0.0001049 allele frequency) (http://exac.broadinstitute.org/ [August 2019]) and also with very low allele frequency in the in-house exome database (0.00045 in > 8000 exomes). The *ASXL1* variant was never observed in ExAc nor in the in-house database. *In silico* predictions for both variants can be seen in Supplementary Table 2.

**Table 2.** Overview of the 21 unique nonsynonymous variants present in both affected family members.

| Gene symbol | Gene ID | Chromosome | Reference | Mutation | Start position | End position | Abberation | Reference Amino Acid | Mutation Amino Acid | Conservation score |
| --- | --- | --- | --- | --- | --- | --- | --- | --- | --- | --- |
| *PDLIM5* | NM_006457 | chr4 | C | T | 95561564 | 95561564 | substitution | R | * | 1.933 |
| *PARP6* | NM_020214 | chr15 | G | A | 72553972 | 72553972 | substitution | H | Y | 9.711 |
| *KTN1* | NM_004986 | chr14 | C | T | 56094742 | 56094742 | substitution | T | I | 3.371 |
| *NEXN* | NM_001172309 | chr1 | A | G | 78399073 | 78399073 | substitution | E | G | 8.789 |
| *PCNX* | NM_014982 | chr14 | C | T | 71428992 | 71428992 | substitution | P | S | 6.163 |
| ***PTPRF*** | **NM_002840** | **chr1** | **A** | **G** | **44072041** | **44072041** | **substitution** | **N** | **S** | **-0.606** |
| *NPR1* | NM_000906 | chr1 | T | C | 153660586 | 153660586 | substitution | L | P | 1.801 |
| *DCAF8* | NM_015726 | chr1 | T | G | 160209533 | 160209533 | substitution | Q | P | 0.727 |
| *C10orf128* | NM_001288742 | chr10 | G | A | 50364232 | 50364232 | substitution | S | L | -0.062 |
| *PYROXD1* | NM_024854 | chr12 | A | G | 21602560 | 21602560 | substitution | K | E | 5.191 |
| *POLR2A* | NM_000937 | chr17 | A | G | 7417024 | 7417024 | substitution | Q | R | 1.397 |
| *CHD3* | NM_001005271 | chr17 | T | C | 7810699 | 7810699 | substitution | L | P | 0.563 |
| *PARD3B* | NM_152526 | chr2 | C | G | 206480223 | 206480223 | substitution | R | G | 3.724 |
| *LANCL1* | NM_001136574 | chr2 | A | C | 211306083 | 211306083 | substitution | F | L | 2.365 |
| ***ASXL1*** | **NM_015338** | **chr20** | **C** | **T** | **31019407** | **31019407** | **substitution** | **R** | **C** | **4.337** |
| *PRMT2* | NM_001242865 | chr21 | C | T | 48071838 | 48071838 | substitution | L | F | -0.331 |
| *GOLIM4* | NM_014498 | chr3 | C | T | 167728086 | 167728086 | substitution | E | K | 3.176 |
| *GTPBP2* | NM_019096 | chr6 | C | A | 43591700 | 43591700 | substitution | E | D | 0.073 |
| *MATN2* | NM_002380 | chr8 | T | C | 98991158 | 98991158 | substitution | C | R | 6.292 |
| *PRUNE2* | NM_015225 | chr9 | G | A | 79267561 | 79267561 | substitution | R | W | 1.445 |
| *AKNA* | NM_030767 | chr9 | C | T | 117139699 | 117139699 | substitution | G | R | -0.126 |

\*stop mutation; conservation score: PhyloP; measures acceleration (faster evolution than expected under neutral drift) as well as conservation (slower than expected evolution). The absolute values of the scores represent $-\log$ p-values under a null hypothesis of neutral evolution.

**Table 3.**
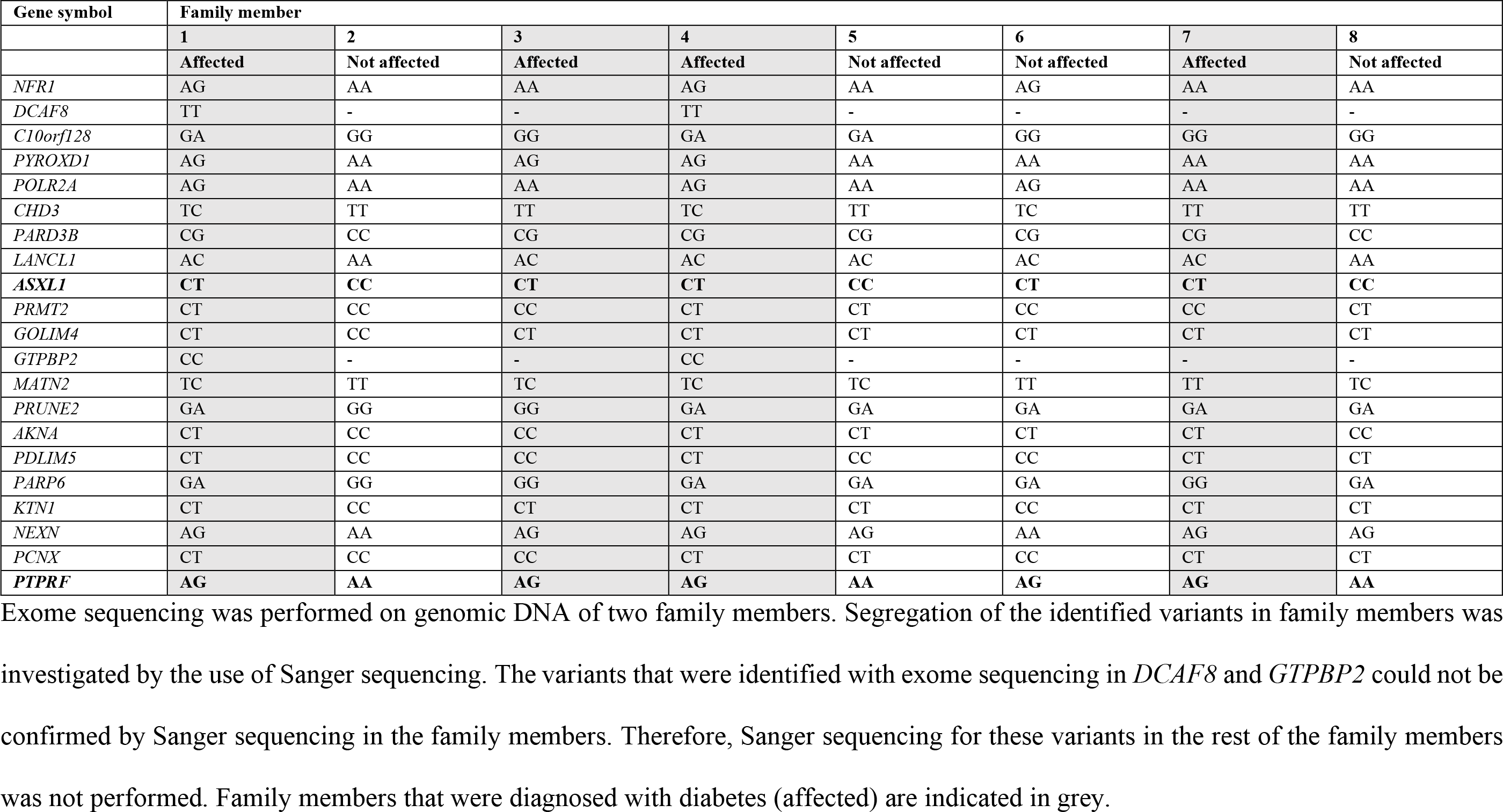
Segregation of the 21 identified variants in all family members.

### PTPRF strongly interacts with proteins involved in insulin signaling

To investigate/identify in which biological processes and pathways the genes *PTPRF* and *ASXL1* are involved, we used the STRING database to search for predicted and published protein-protein interactions. Analysis showed that *PTPRF* strongly interacts with several members of the insulin signaling pathway. High confidence scores were assigned to the interactions between *PTPRF* and *Insulin receptor (INSR)* and *Insulin receptor substrate* 1 and *2 (IRS1* and 2) (Table 4, Figure 1A). Indeed, *PTPRF* was demonstrated to be expressed in several insulin sensitive tissues where it interacts with the insulin receptor and dephosphorylates its tyrosine-kinase domain, thereby inhibiting its activation and downstream signaling [20]. These results point out to an important role for *PTPRF* in insulin signaling. Protein-protein interaction analysis also revealed that *ASXL1* did not interact with proteins involved in insulin signaling, glucose metabolism or other biological processes that are likely to play a role in the pathogenesis of type 2 diabetes (Table 4 and Figure 1B).

**Table 4.**
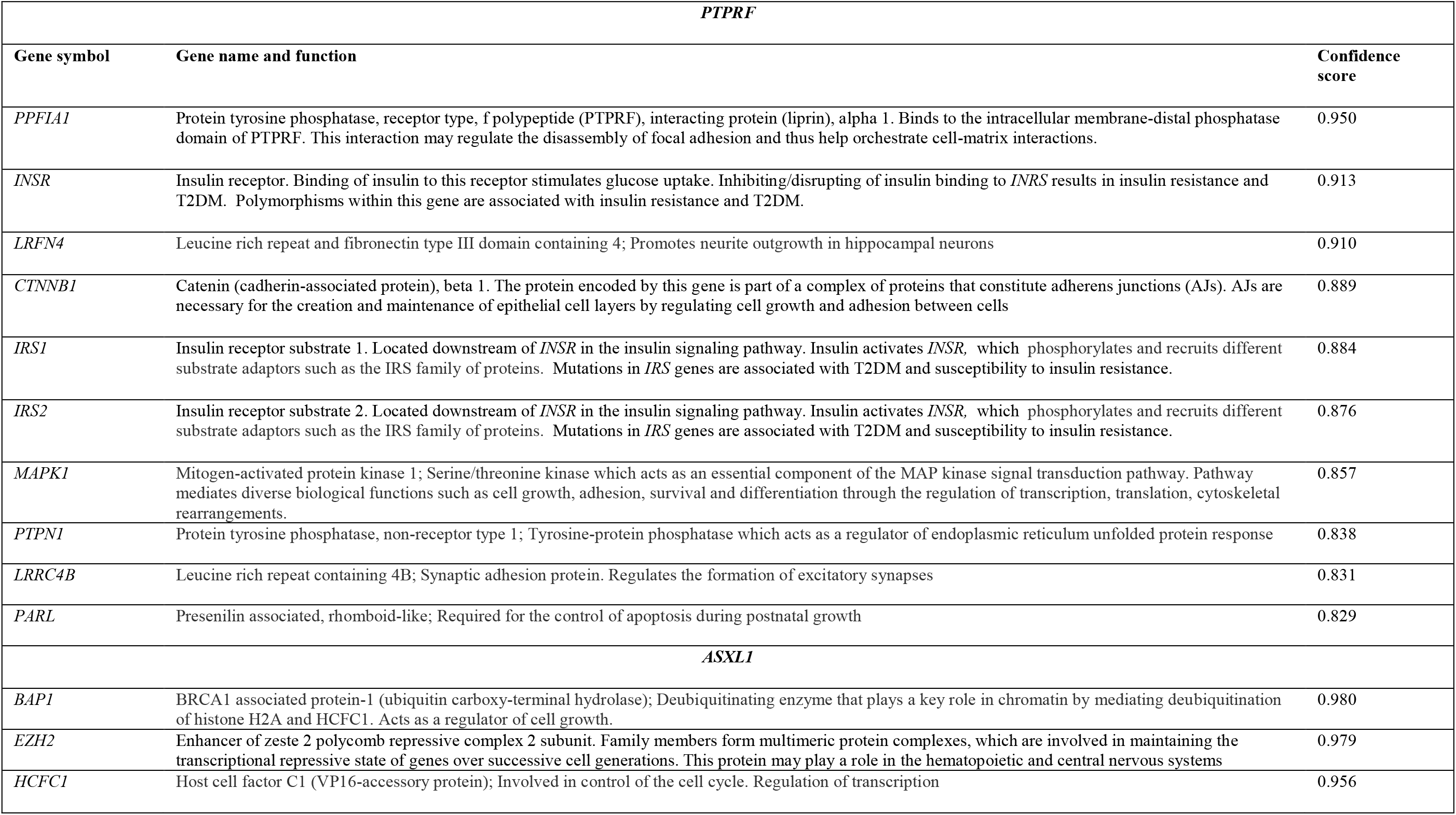
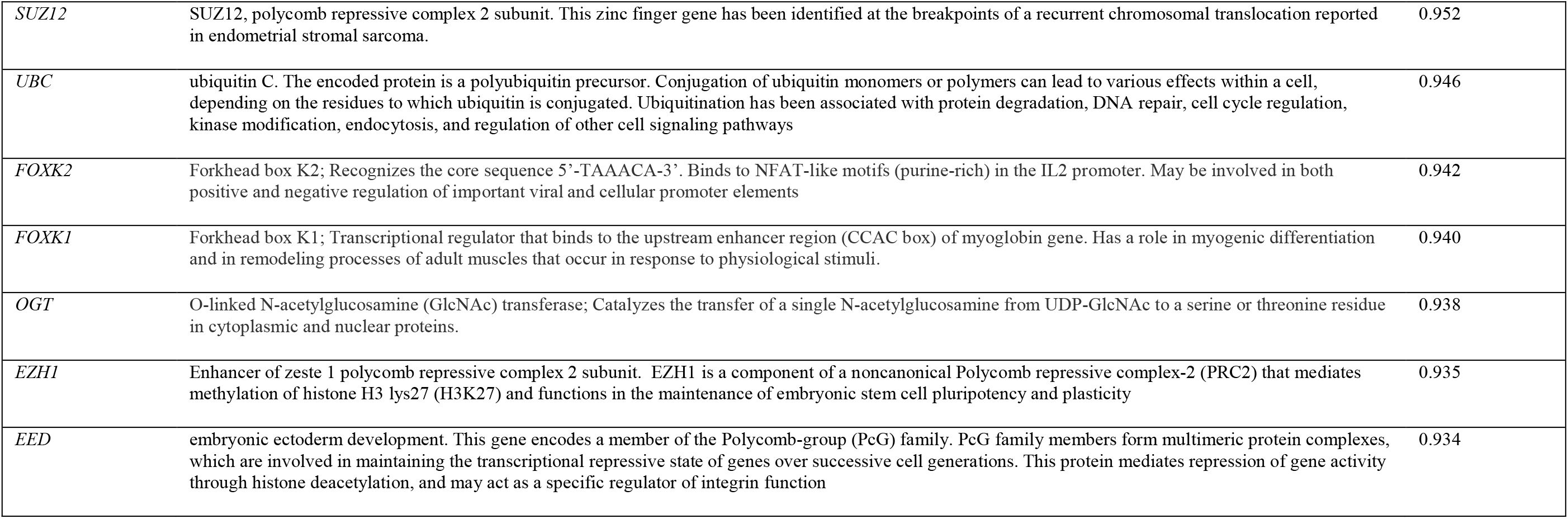
Predicted interaction partners for *PTPRF* and *ASXL1*.

| *PTPRF* |  |  |
| --- | --- | --- |
| Gene symbol | Gene name and function | Confidence score |
| *PPFIA1* | Protein tyrosine phosphatase, receptor type, f polypeptide (PTPRF), interacting protein (liprin), alpha 1. Binds to the intracellular membrane-distal phosphatase domain of PTPRF. This interaction may regulate the disassembly of focal adhesion and thus help orchestrate cell-matrix interactions. | 0.950 |
| *INSR* | Insulin receptor. Binding of insulin to this receptor stimulates glucose uptake. Inhibiting/disrupting of insulin binding to *INSR* results in insulin resistance and T2DM. Polymorphisms within this gene are associated with insulin resistance and T2DM. | 0.913 |
| *LRFN4* | Leucine rich repeat and fibronectin type III domain containing 4; Promotes neurite outgrowth in hippocampal neurons | 0.910 |
| *CTNNB1* | Catenin (cadherin-associated protein), beta 1. The protein encoded by this gene is part of a complex of proteins that constitute adherens junctions (AJs). AJs are necessary for the creation and maintenance of epithelial cell layers by regulating cell growth and adhesion between cells | 0.889 |
| *IRS1* | Insulin receptor substrate 1. Located downstream of *INSR* in the insulin signaling pathway. Insulin activates *INSR*, which phosphorylates and recruits different substrate adaptors such as the IRS family of proteins. Mutations in *IRS* genes are associated with T2DM and susceptibility to insulin resistance. | 0.884 |
| *IRS2* | Insulin receptor substrate 2. Located downstream of *INSR* in the insulin signaling pathway. Insulin activates *INSR*, which phosphorylates and recruits different substrate adaptors such as the IRS family of proteins. Mutations in *IRS* genes are associated with T2DM and susceptibility to insulin resistance. | 0.876 |
| *MAPK1* | Mitogen-activated protein kinase 1; Serine/threonine kinase which acts as an essential component of the MAP kinase signal transduction pathway. Pathway mediates diverse biological functions such as cell growth, adhesion, survival and differentiation through the regulation of transcription, translation, cytoskeletal rearrangements. | 0.857 |
| *PTPN1* | Protein tyrosine phosphatase, non-receptor type 1; Tyrosine-protein phosphatase which acts as a regulator of endoplasmic reticulum unfolded protein response | 0.838 |
| *LRRC4B* | Leucine rich repeat containing 4B; Synaptic adhesion protein. Regulates the formation of excitatory synapses | 0.831 |
| *PARL* | Presenilin associated, rhomboid-like; Required for the control of apoptosis during postnatal growth | 0.829 |
| *ASXL1* |  |  |
| *BAP1* | BRCA1 associated protein-1 (ubiquitin carboxy-terminal hydrolase); Deubiquitinating enzyme that plays a key role in chromatin by mediating deubiquitination of histone H2A and HCFC1. Acts as a regulator of cell growth. | 0.980 |
| *EZH2* | Enhancer of zeste 2 polycomb repressive complex 2 subunit. Family members form multimeric protein complexes, which are involved in maintaining the transcriptional repressive state of genes over successive cell generations. This protein may play a role in the hematopoietic and central nervous systems | 0.979 |
| *HCFC1* | Host cell factor C1 (VP16-accessory protein); Involved in control of the cell cycle. Regulation of transcription | 0.956 |
| *SUZ12* | SUZ12, polycomb repressive complex 2 subunit. This zinc finger gene has been identified at the breakpoints of a recurrent chromosomal translocation reported in endometrial stromal sarcoma. | 0.952 |
| *UBC* | ubiquitin C. The encoded protein is a polyubiquitin precursor. Conjugation of ubiquitin monomers or polymers can lead to various effects within a cell, depending on the residues to which ubiquitin is conjugated. Ubiquitination has been associated with protein degradation, DNA repair, cell cycle regulation, kinase modification, endocytosis, and regulation of other cell signaling pathways | 0.946 |
| *FOXK2* | Forkhead box K2; Recognizes the core sequence 5'-TAAACA-3'. Binds to NFAT-like motifs (purine-rich) in the IL2 promoter. May be involved in both positive and negative regulation of important viral and cellular promoter elements | 0.942 |
| *FOXK1* | Forkhead box K1; Transcriptional regulator that binds to the upstream enhancer region (CCAC box) of myoglobin gene. Has a role in myogenic differentiation and in remodeling processes of adult muscles that occur in response to physiological stimuli. | 0.940 |
| *OGT* | O-linked N-acetylglucosamine (GlcNAc) transferase; Catalyzes the transfer of a single N-acetylglucosamine from UDP-GlcNAc to a serine or threonine residue in cytoplasmic and nuclear proteins. | 0.938 |
| *EZH1* | Enhancer of zeste 1 polycomb repressive complex 2 subunit. EZH1 is a component of a noncanonical Polycomb repressive complex-2 (PRC2) that mediates methylation of histone H3 lys27 (H3K27) and functions in the maintenance of embryonic stem cell pluripotency and plasticity | 0.935 |
| *EED* | embryonic ectoderm development. This gene encodes a member of the Polycomb-group (PcG) family. PcG family members form multimeric protein complexes, which are involved in maintaining the transcriptional repressive state of genes over successive cell generations. This protein mediates repression of gene activity through histone deacetylation, and may act as a specific regulator of integrin function | 0.934 |

**Figure 1.**
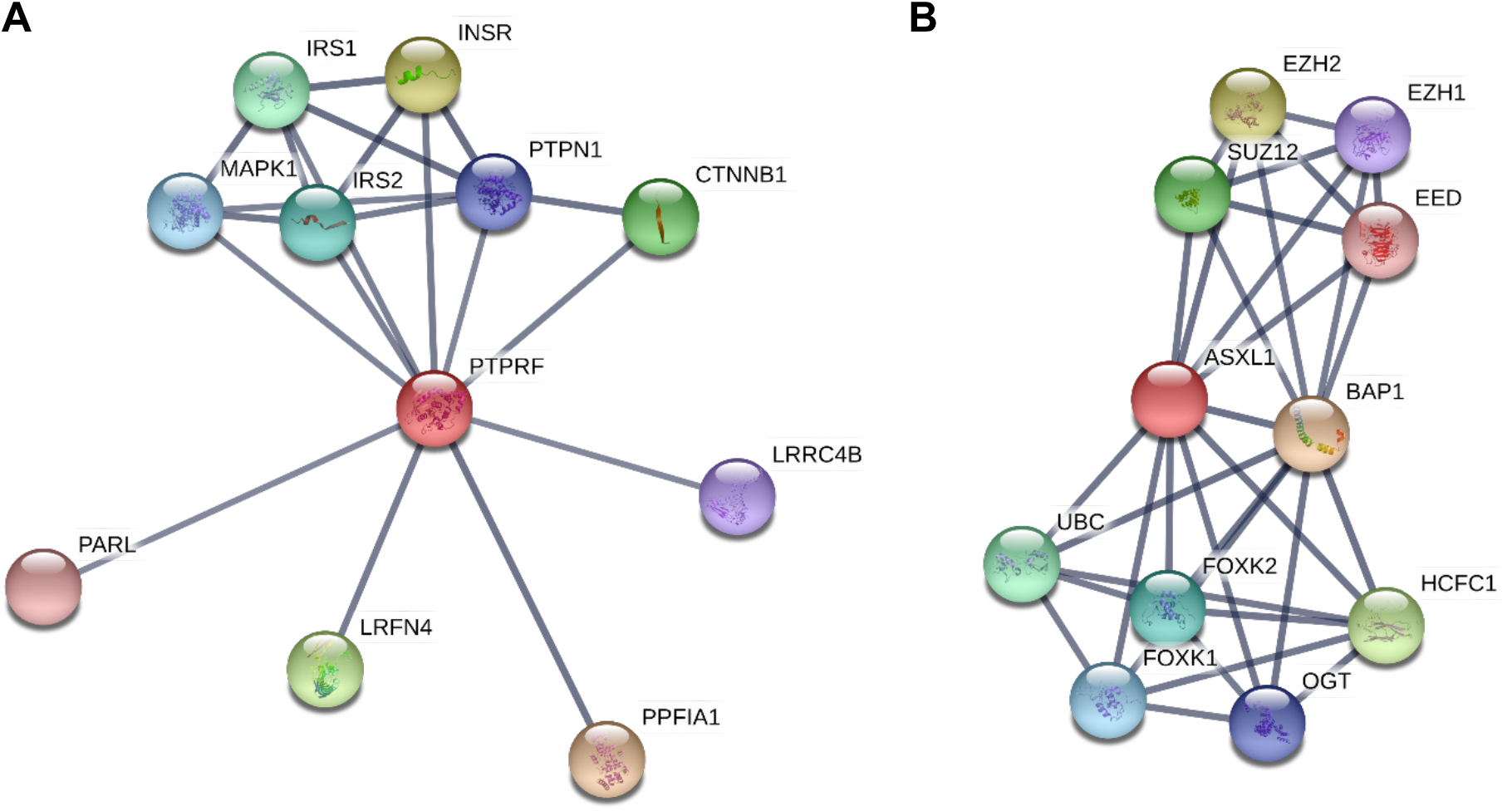
Protein-protein interaction (PPI) networks of *PTPRF* and *ASXL1*. PPI networks of **(A)** *PTPRF* and **(B)** *ASXL1*. A confidence score is assigned to each interaction based upon direct (physical) and indirect (functional) associations. Confidence score cut-off was ≥0,7 (high confidence)

### Investigation of the PTPRF gene in an additional cohort of T2DM patients

The results described above clearly pointed out to *PTPRF* as a strong candidate gene for this T2DM phenotype. To validate our results, we investigated the *PTPRF* gene for the possible presence of genetic variants in an additional cohort of 24 unrelated T2DM patients. These patients were selected based upon disease characteristics that matched the characteristics of the affected family members (insulin > 1U kg or 100 U/day; TG > 1,5 mmol/l; BMI < 35 kg m^2^). All coding exons of the *PTPRF* gene were sequenced in these 24 patients by the use of Ion torrent sequencing. The c.3614A>G variant was not present in these 24 patients. Also no additional rare variants were detected in this cohort.

## Discussion

In the present study, we applied whole-exome sequencing to identify genetic variants possibly involved in disease susceptibility in a family with T2DM, characterized by substantial insulin resistance, hypertension and hypertriglyceridemia in the absence of hypercholesterolemia. We identified the rare variant c.3614>G, located within the *PTPRF* gene, as a novel candidate susceptibility variant. Sequencing of all coding exons of the *PTPRF* gene in a cohort of 24 non-related T2DM patients did not reveal any additional variants. Our results suggest that the c.3614A>G variant might play a role in an unusual T2DM subtype that is characterized by a severe insulin resistance and “isolated” hypertriglyceridemia. Identification of genetic variants responsible for specific forms of diabetes provide new insights regarding underlying disease aetiology. These new insights may help to improve diagnosis and to optimize treatment strategies in these specific T2DM subtypes.

The c.3614A>G variant was detected in all affected family members but also in one unaffected family member. Although this family member was not diagnosed with type 2 diabetes, his triglyceride levels were elevated. All family members with diabetes had episodes with grossly elevated TG levels (>10 mmol/L), fairly independent of age, but occasionally also showed less elevated TG levels. TG level was, however, always above the normal range. TG level in the genetically affected family member without diabetes was moderately elevated in one single blood sample. Perhaps this person had a combination of relative active lifestyle and excellent beta-cell function, resulting in absence of diabetes. Additionally, T2DM is a multifactorial disease in which both genetic as environmental factors play a role. Another reason why this individual carries the c.3614A>G variant but was not diagnosed with T2DM might be reduced penetrance. As often seen for multifactorial diseases with a heritable component, the c.3614A>G variant may have reduced penetrance, meaning that some individuals fail to express the trait, even though they carry the allele. The degree of penetrance is difficult to determine and is often age related. Given that the onset of symptoms in T2DM are age related and affected by both genetic as well as environmental factors such as nutrition, it is possible that this individual will develop the disease in the near future, although he is currently older than the age at which the diabetes was diagnosed in his brothers.

Although we cannot exclude the variant located in *ASXL1* as a candidate, the evidence for *PTPRF* as a candidate gene for T2DM is more convincing. Our finding that a genetic variant within *PTPRF* may be involved in the pathogenesis of type 2 diabetes is supported by earlier studies. Miscio and co-workers investigated whether common polymorphisms within the *PTPRF* gene were associated with insulin resistance. They identified a single nucleotide polymorphism (SNP) within the promoter region (−127T>A; rs3001722) in which the A allele (minor allele) was associated with lower body mass index (BMI), waist circumference and mean blood pressure in a cohort of 589 unrelated non-diabetic subjects. In addition, the A allele was associated with lower triglycerides, glucose and insulin levels during an oral glucose tolerance test (oGTT) in an independent cohort of 307 unrelated non-diabetic individuals [21]. Menzaghi and co-workers report an association between the SNP 2518A>G (rs2782641), located within intron 3, and cononary artery disease (CAD) in a T2DM patient cohort including 592 individuals [22]. To our knowledge, the c.3614A>G variant has not been reported in literature in relation to T2DM. These studies, together with our results, suggest that genetic variants within *PTPRF* are contributing to insulin resistance and T2DM in at least a subpopulation of patients.

*PTPRF* is a receptor-type transmembrane protein tyrosine phosphatase (PTPase) that is expressed in several insulin target tissues [23]. It physically interacts with the insulin receptor and dephosphorylates its tyrosine-kinase domain thereby inhibiting insulin signaling [24]. Transgenic mice overexpressing *PTPRF* are insulin resistant [25] and, conversely, *PTPRF* knock-out mice are characterized by increased insulin sensitivity [26]. *PTPRF* is therefore a potential candidate gene for insulin resistance. Regarding *ASXL1*, no studies were reported showing evidence that *ASXL1* is involved in the pathogenesis of T2DM.

Most studies investigating the genetic background of T2DM are focusing on identifying common variants that are associated with the disease [8, 27-29]. However, most studies have not been particularly successful in discovering new disease genes. As a consequence, the rare variant hypothesis [30, 31], which states that rare variants with large effects are the primary drivers of common diseases, has received renewed attention. Studies investigating Mendelian diseases have demonstrated that a family-based approach is a good method to detect these rare variants with relatively high impact on disease [32]. Rare variants are not present in the general population at any reasonable frequency and an advantage of a family-based study is that more observations of a rare variant can be made in large families. However, type 2 diabetes is a late-onset disease and recruiting such families is challenging. We identified this family because of the unusual phenotype of the index patient, and noted the familial existence of the combination of severe insulin resistance and “isolated” hypertriglyceridemia: hypertriglyceridemia in the absence of hypercholesterolemia. As expected, hypertriglyceridemia was associated with decreased HDL-cholesterol levels. In this study, we used the family-based approach combined with whole-exome sequencing to detect variants that may have a role in susceptibility for the disease in this family. We were able to identify a rare variant located in a strong T2DM candidate gene, which suggests that the family-based approach combined with whole-exome sequencing may be a useful strategy to investigate the genetic background of type 2 diabetes.

While our findings suggest that the c.3614A>G variant in *PTPRF* may underly the phenotype of the this family, we were unable to identify a second family within our facility that resembles the disease characteristics observed in this first family, despite sharing this as a candidate gene via genematcher.com [33, 34]. In addition, no variants in *PTPRF* were identified in a patient cohort of 24 unrelated T2DM patients who phenotypically matched the characteristics of the affected family members. Our study lacks confirmation through functional studies, which should be included in future investigations. We realize that it is possible that the identified variant is a false positive result. However, we would like to stress that the variant in *PTPRF* was identified using an unbiased, non-candidate gene driven, whole exome-wide approach and that the gene is known to be involved in insulin signaling and associated with metabolic disturbances. To our opinion, this suggests that *PTPRF* is a potential candidate gene that, at least, should be added to the discussion regarding genetic variants underlying T2DM. In that light, it might be best to see this report as a case report providing researchers and clinicians the opportunity to respond and set up collaborations in order to identify additional cases/families.

In summary, we applied whole-exome sequencing to identify genetic variants possibly involved in T2DM disease susceptibility in a family with highly aggregated type 2 diabetes. Our results suggest that the c.3614A>G variant in the *PTPRF* gene play a role in an unusual T2DM subtype that is characterized by a severe insulin resistance and isolated hypertriglyceridemia. These results indicate that combining the family-based approach with whole-exome sequencing may be a useful strategy to further unravel the genetic background of type 2 diabetes. The genetic variant identified within this study represents a promising candidate for stratification of T2DM patients regarding mild or severe insulin resistance. Follow-up studies are needed to confirm that the identified variant in the *PTPRF* gene is indeed responsible for severe insulin resistance and isolated hypertriglyceridemia and to further unravel the precise working mechanism responsible for this unusual T2DM phenotype. If several independent studies are able to confirm our initial results, this genetic variant may have additional value for diagnosis and to optimize treatment strategies in patients with this unusual T2DM subtype that is characterized by severe insulin resistance and isolated hypertriglyceridemia.

## Data Availability

Availability of data and material
Please contact author for data requests

**List of abbreviations**
ALAT: alanine aminotransferase
ASAT: Aspartaat-aminotransferase
BMI: body mass index
CAD: coronary artery disease
GP: general practitioner
GWAS: genome-wide association study
HbA_1c_: hemoglobin A1c
HDL: high-density lipoprotein
LDL: low-density lipoprotein
oGTT: oral glucose tolerance test
PCR: polymerase chain reaction
protein-protein: interaction
SNP: single nucleotide polymorphism
SU: Sulphonylureas
T2DM: Type 2 diabetes mellitus
TG: triglycerides

## Acknowledgements

The authors would like to thank Lisenka E.L.M. Vissers, Petra de Vries, Marloes Steehouwer and Christian Gilissen for their help and advice during this study. The authors are very grateful to the patients and family members who made it possible to perform this study.

## Funding

E.T. was supported by an EFSD/Novo Nordisk research grant provided by the European Foundation for the Study of Diabetes (EFSD) and by a grant of the Else-Kröner-Fresenius-Stiftung.

## Conflicts of interests

None declared

## Declarations

### Ethics approval and consent to participate

The authors state that they have obtained appropriate institutional review board approval or have followed the principles outlined in the Declaration of Helsinki for all human or animal experimental investigations. All family members were informed about the goal of the study and consented to participation. Study was approves by the The local Medical Ethical Committee, Commissie Mensgebonden Onderzoek (CMO) regio Arnhem-Nijmegen.

## Consent for publication

Consent to publish was obtained from all individuals.

### Availability of data and material

Please contact author for data requests

## Author contributions

ET: designed the study, performed the experiments, analyzed and interpreted the results, wrote the manuscript. MK: performed the experiments, analyzed and interpreted the results. AH: designed the study, analyzed and interpreted the results, revised the manuscript. CT: designed the study, analyzed and interpreted the results, revised the manuscript.

## Notes

### Competing Interest Statement

The authors have declared no competing interest.

### Clinical Trial

Ethics approval and consent to participate
The authors state that they have obtained appropriate institutional review board approval or have followed the principles outlined in the Declaration of Helsinki for all human or animal experimental investigations. All family members were informed about the goal of the study and consented to participation. Study was approves by the The local Medical Ethical Committee, Commissie Mensgebonden Onderzoek (CMO) regio Arnhem-Nijmegen.
Consent for publication
Consent to publish was obtained from all individuals.

### Author Declarations

Study was approves by the The local Medical Ethical Committee, Commissie Mensgebonden Onderzoek (CMO) regio Arnhem-Nijmegen.

